# Identifying potential effects of relative age in school year: an instrumental variable phenome-wide association study in the UK Biobank

**DOI:** 10.1101/2023.05.26.23290586

**Authors:** Melanie A de Lange, Neil M Davies, Louise AC Millard, Kate Tilling

## Abstract

**Background:** Observational research shows that a child’s relative age within their school year (‘relative age’) is associated with educational attainment and mental health. However, previous studies have only examined a small number of outcomes and evidence of the persistence of effects into adulthood is mixed. We conducted a hypothesis-free investigation of the effects of relative age.

**Method:** We used a regression discontinuity design and an instrumental variable (IV)-pheWAS in the UK Biobank (participants aged 40-69 years at baseline), using the PHESANT software package. We created two IVs for relative age: being born in September vs. August (*n*=64 075) and week of birth (*n*=383 309). Outcomes passing the Bonferroni-corrected *P* value threshold for either instrument were plotted to identify those displaying a discontinuity at the school year transition.

**Results:** We found 21 traits associated with at least one of the instruments (*P* value below the Bonferroni threshold). Of these, 13 showed a discontinuity at the school year transition. These included previously identified effects including those with a younger relative age being less likely to have educational qualifications and more likely to have started smoking at an earlier age. We also identified a novel potential effect of a younger relative age in school year causing a better lung function as adults.

**Conclusion:** Educational policy should address educational inequality due to relative age. Further research should seek to replicate our identified effect on lung function in different populations, and investigate the mechanisms through which this effect may act.

**Key Messages:**

- Children’s relative age within their school year has been associated with mental health in childhood and educational attainment.
- Our results supported previously identified effects, with those who were younger in their school year being less likely to have educational qualifications and more likely to report starting smoking at an earlier age.
- We also found a potential beneficial effect of a younger relative age in school year on lung function in adulthood.

## Introduction

Children’s age within their school year, henceforth referred to as relative age, consistently relates to educational attainment. Children who are born later in the school year have lower average educational attainment, when it comes to tests taken on the same date, than those born at the beginning of the school year^1-6^. This means that in England, where entry for each school year runs from the 1st September to the 31st August, summer-born children on average perform worse than autumn-born children^7^.

Relative age differences in educational attainment occur across a range of education systems with different school year start dates, so are unlikely to be due to season of birth^8^. Although educational differences are greatest when comparing those born at the start and end of the school year (e.g. September vs. August in England), there is evidence of a linear change in educational outcomes over relative age^7^. These differences are largest when children start school and do diminish with age, but are still evident when children finish compulsory schooling^7^.

Children born later in the school year are also more likely to be diagnosed with special educational needs^9-14^, be bullied at school ^10, 15^, engage in risky behaviour such as underage smoking^7^, and commit a crime in adolescence^16^. They also tend to have poorer mental health during childhood^14, 17-19^ and higher suicide rates in adolescence and young adulthood (age 15-25)^20^.

A limited number of studies have examined whether the effects of relative age persist into adulthood. Some studies have found negative effects on whether adults are employed^21^, as well as their earnings^22-24^, self-confidence and risk taking^25^. Others have found little effect on whether people are in work and earnings^7, 21, 26, 27^, or health and mental wellbeing during adulthood^7, 19, 21^.

Many studies of relative age have used a regression discontinuity design (RDD). This is a quasi-experimental method that can be used be used to examine the effects of policies which assign people to a treatment or intervention based on a threshold on a continuous variable^28, 29^. Because a child’s birth date cannot be controlled precisely (assuming natural birth), whether a child is assigned to the group born before or after the school year entry cut-off is quasi-random^19^. This means that, close to the threshold, covariates should be balanced between those born just before or just after the cut-off^19^. Thus, the RDD approach can give unbiased estimates of causal effects if the window around the threshold is small, and the underlying continuous variable (here the date of birth) cannot be manipulated ^28^.

To date, research investigating relative age effects has focused on a small number of outcomes, mostly educational or related to mental health and wellbeing. In contrast to such hypothesis-driven approaches, hypothesis-free approaches test all measured factors, regardless of previous knowledge^30^. Phenome scans are a type of hypothesis-free analysis which can identify novel associations by testing the association of a trait of interest with a large number of other phenotypes (“the phenome”)^31^. PHESANT is a software package for performing comprehensive phenome scans in UK Biobank^31^. Rather than being limited to a homogenous subset of phenotypes that can be analysed in the same way, PHESANT enables users to scan a heterogenous set of phenotypes (all UK Biobank continuous, integer and categorical fields)^31^. An instrumental variable (IV)-pheWAS is a type of pheWAS that uses an instrumental variable in order to investigate causality.

In this study we used PHESANT to conduct two IV-pheWAS, using a regression discontinuity, to identify phenotypes associated with relative age within ∼383 000 UK Biobank participants born in England.

## Method

### Study population

The UK Biobank is a prospective cohort of around 500 000 adults aged mainly 40-69 years when recruited in in 2006-2010^32^ (5.5% participation rate^33^). It contains extensive phenotypic data collected via lifestyle and health questionnaires; physical assessments; and blood, saliva and urine samples^32^. This research was conducted under UK Biobank application 16729 (dataset ID 48196).

Of the 502 448 UK Biobank participants, we removed 111 994 participants who were not born in England (as other UK countries have different school entry cut-off dates). We also removed 27 participants who withdrew their consent, giving a sample of 390 427 (see Figure 1).

**Figure 1.**
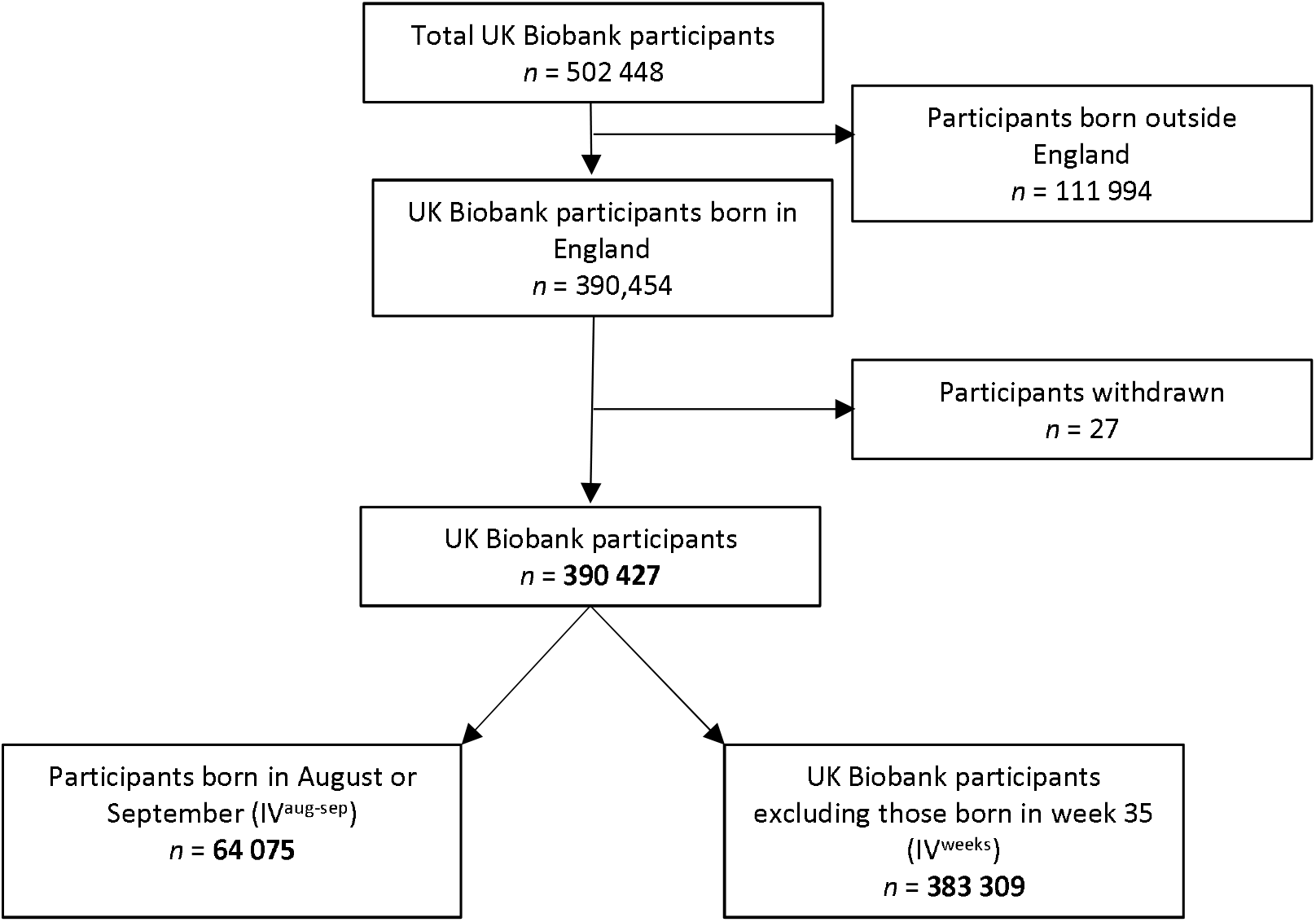
Participant flow diagram

### Instrumental variables (IVs) for relative age

Month of birth and week of birth were acquired from a central registry and updated by participants at their initial UK Biobank assessment centre visit. These variables were used to create two IVs for relative age. IV^sep-aug^ was a binary variable restricted to participants born in September or August, with September coded as one and August as zero. IV^weeks^ was week of birth as a continuous variable, with the first full week in September coded as zero and the last full week of August coded as 51. We excluded the week containing the August – September crossover in order to avoid misclassification (as we cannot know if these participants were born in August or September).

### Covariates

We included age at recruitment and sex as covariates in our models to reduce variation in our outcomes and increase precision. Age was derived from date of birth and date of attending initial assessment, and provided to us from UK Biobank as an integer. Sex was acquired from the NHS central registry at recruitment but was updated by the participant in some cases.

### Deriving outcomes with PHESANT

The UK Biobank Showcase (https://biobank.ndph.ox.ac.uk/showcase/) provides a searchable directory of variables available in the UK Biobank based on field type (integer, continuous, categorical (single) and categorical (multiple)). PHESANT was run with the ‘save’ option on to derive cleaned variables to be used as outcomes in the IV-pheWAS. This involved PHESANT categorising each outcome variable into one of four data types: continuous, ordered categorical, unordered categorical or binary, using its automated rule-based method described elsewhere^31^. The derived outcomes were stored as CSV files labelled according to data type.

### Statistical methods

#### IV-pheWAS

We tested the association between our two IVs and our derived outcome variables. Regression analyses were adjusted for age and sex. We only tested outcomes with at least 500 participants, and at least 10 participants in each category in binary and unordered categorical variables. *P* values were generated for each outcome, using a likelihood ratio test comparing the model including the IV as a covariable versus the same model but not including the IV as a covariable. Results were ranked according to *P* value. In studies that test multiple hypotheses simultaneously, the family-wise error rate (the chances of getting type one errors (false positives)) rises as the number of tests increases^34^. To take multiple testing into account when evaluating the strength of the evidence of our results, we defined potential effects as any association of relative age with an outcome that passed the stringent Bonferroni-corrected significance threshold (0.05/number of tests performed) in either IV-pheWAS. This controls the family-wise error rate assuming that tests are independent^35^.

We also ran a third IV-pheWAS, IV-months, where our IV was a set of 11 indicator variables describing the month of birth of each participant (excluding September) and compared each of the 11 months to September. However, we have not included this in our main results as it picked up many associations that were not specific to the discontinuity we were looking for (see Table S1 for results).

To explore whether our identified potential effects displayed a discontinuity at the transition between school years, and therefore were relative age effects rather than seasonal trends, estimates and confidence intervals for IV^months^ (month of birth relative to September) were plotted for each potential effect.

### Sensitivity Analyses

The results of IV^months^ were used to run a meta-regression analyses for each identified potential effect. For each potential effect we ran 11 meta-regressions, placing the origin of the linear function at the start of each month from October to August. We identified the model with the largest association. If the this model was for the time-origin of October, this was taken as evidence of a relative age in school year effect.

Analyses were performed in R version 4.1.0 and Stata version 17. Code is available at https://github.com/MRCIEU/PHESANT-IV-pheWAS-relative-school-age (git tag v0.2 corresponds to the version presented here).

## Results

### IV-pheWAS results

Out of the 15 034 tests performed for IV^sep-aug^ and 23 164 tests performed for IV^weeks^, we found 21 traits that had a *P* value below the Bonferroni threshold in at least one of these IV-pheWAS (Table 1; See Table S1 for full results for both IVs (note we do not have estimates for unordered categorical variables)). The eight traits associated with IV^sep-aug^ were also associated with IV^weeks^. Model convergence was not reached for a minority of outcomes and as a result these outcomes were not considered further (See Table S2 for a list of these variables).

**Table 1.**
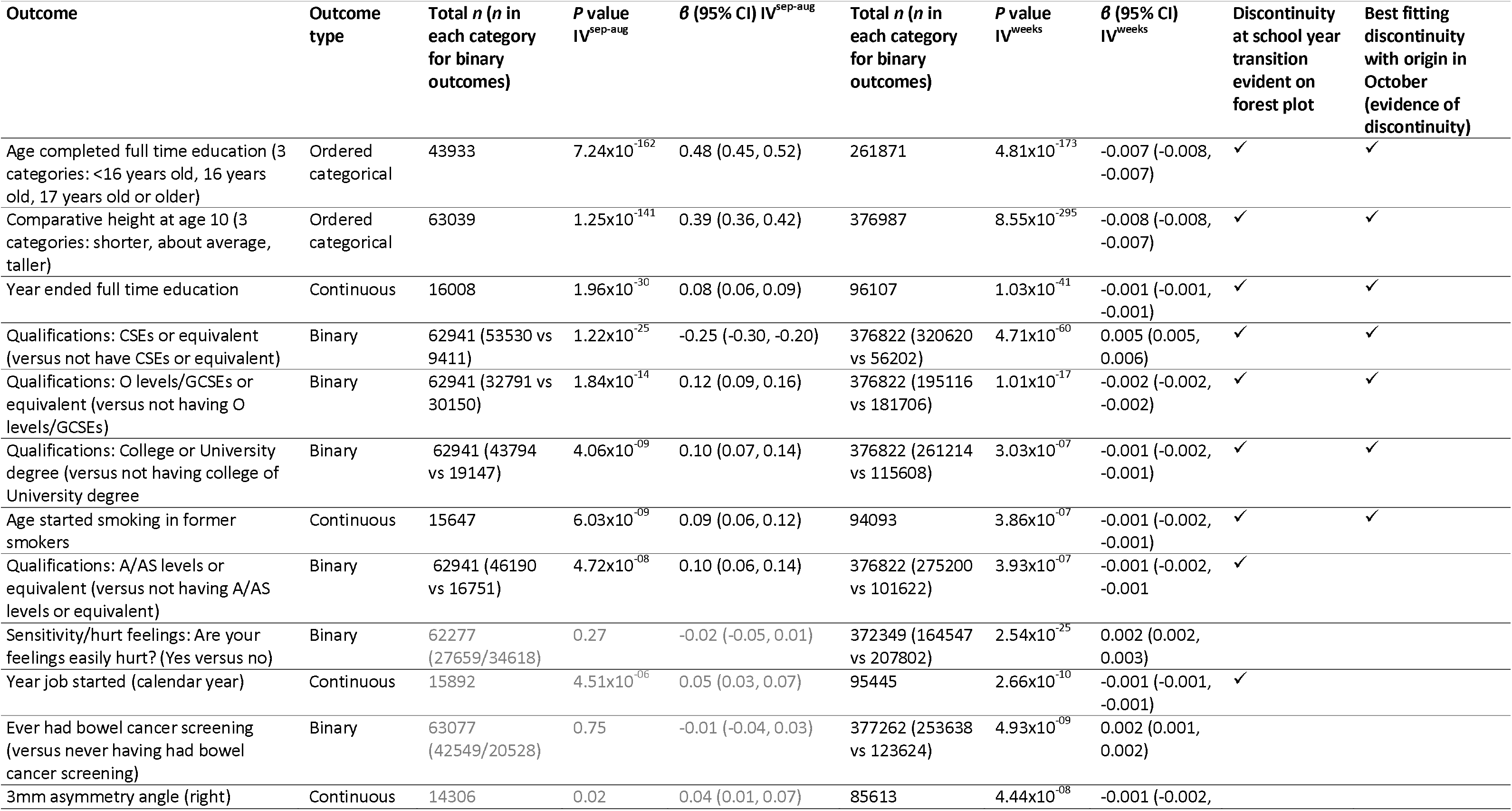
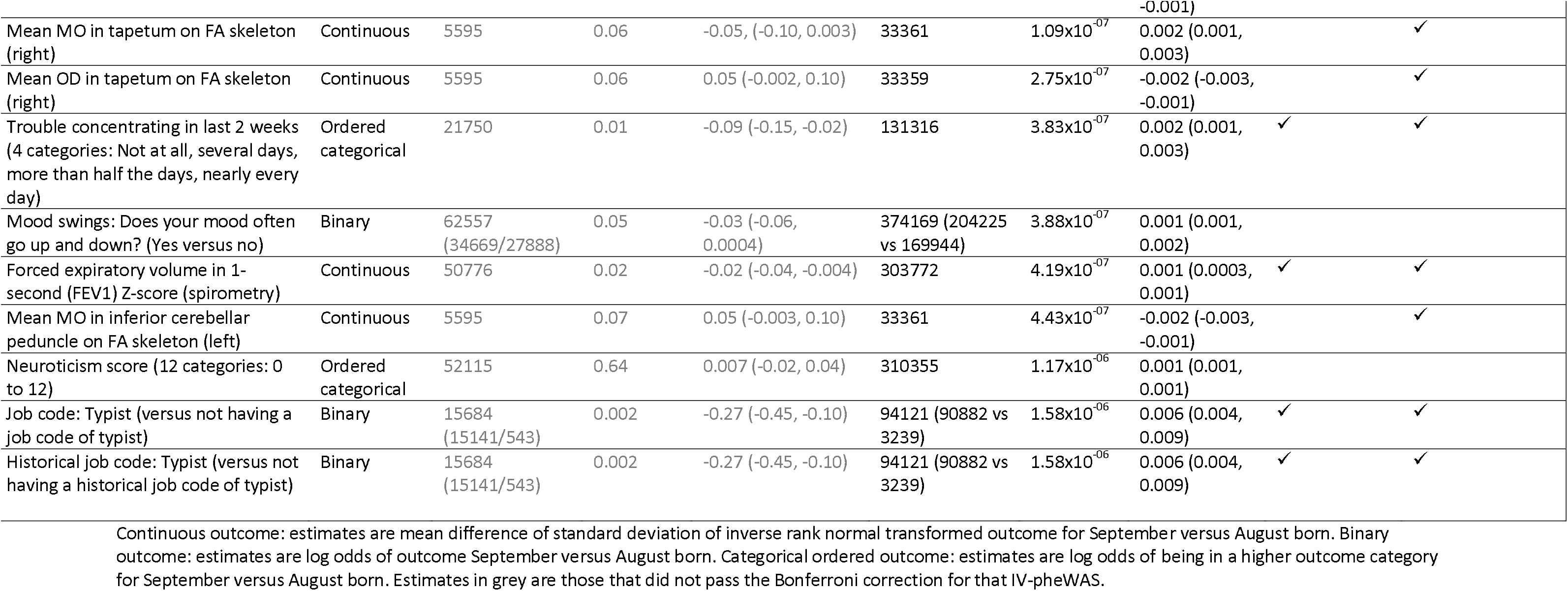
Traits associated with being born in September vs August (IV^sep-aug^) or week of birth (IV^weeks^)

Six of the eight traits that passed the Bonferroni-corrected threshold (*P*=3.33x10^−6^) for IV^sep-aug^ were related to education (see Table 1). For example, participants born in September were more likely to have O levels/GCSEs (*P*=1.84x10^−14^), A/AS levels (*P*=4.72x10^−08^) or a college/university degree (*P*=4.06x10^−09^) and less likely to have CSEs (*P*=1.22x10^−25^) than those born in August. Those born in September were also more likely to complete full-time education at an older age than those born in August (*P*=7.24x10^−162^). These findings are supported by the results of IV^weeks^ which found that participants born in a later week in the academic year had worse education outcomes (see Table 1).

Moving beyond education, participants born in September vs. August, and participants with an earlier week of birth in school year, were more likely to report being comparatively taller for their age at age 10 (*P*=1.25x10^−141^ and *P*=8.55x10^−295^). Participants born in September vs. August, and participants with an earlier week of birth in school year, who are former smokers also report starting smoking later (*P*=6.03x10^−09^ and 3.86x10^−07^). However, little evidence was seen amongst current smokers (*P*=0.32 and *P*=0.16), although this may be due to the smaller sample size in this group hence lower statistical power.

A further 13 traits passed the Bonferroni-corrected threshold (*P*=2.16x10^−6^) for IV^weeks^ alone. In terms of psychological traits, IV^weeks^ was positively associated with having your feelings hurt easily (*P*=2.54x10^−25^), having mood swings (*P*=3.88x10^−07^) and participants’ neuroticism scores (*P*=1.17x10^−06^), meaning those with a younger relative age (later week of birth) were more likely to have these traits. Week of birth was also found to be associated with a number of physical measurements, including being positively associated with forced expiratory volume in one second z-score (4.19x10^−07^). IV^weeks^ was also negatively associated with 3mm asymmetry angle (right eye) (*P*=4.44x10^−08^), mean OD in tapetum on FA skeleton (right) (*P*=2.75x10^−07^) and mean MO in inferior cerebellar peduncle on FA skeleton (left) (*P*=4.43x10^−07^), and positively associated with mean MO in tapetum on FA skeleton (right) (1.09x10^−07^). However, the fact that brain and skeleton MRI, and eye measurement phenotypes passed the Bonferroni threshold may be the result of the sheer number of these variables in the UK Biobank, rather than true effects.

IV^weeks^ was positively associated with ever being screened for bowel cancer (*P*=4.93x10^−09^) but little evidence of an association with people reporting having had bowel cancer itself (small bowel cancer *P*=0.89; large bowel cancer *P*=0.19), possibly due to inadequate statistical power. IV^weeks^ was positively associated with having a job code or historical job code of typist (*P*=1.58x10^−06^ for both). It was also positively associated with answering yes to the question “Over the last 2 weeks, how often have you been bothered by trouble concentrating on things, such as reading the newspaper or watching television?” (*P*=3.83x10^−07^). Another related phenotype, “Over the last 2 weeks, how often have you been bothered by any of the following problems? [depressive symptoms] Trouble concentrating on things, such as reading the newspaper or watching television”, was associated in a direction consistent with our finding (*P*=0.03), although it did not pass the Bonferroni threshold.

### Meta-analysis forest plots

Of the 21 potential effects identified in the IV-pheWAS, 20 had an output from PHESANT for IV^months^ and were plotted by month of birth. A discontinuity at the transition point between school years was visually identified for 13 of these (see Table 1 and Figures S1-S20) – educational qualifications (CSEs, O level/GCSEs, A/AS Levels, College/University degree), the year and age full time education ended, the year a person’s job started, comparative height at age 10, the age former smokers started smoking, having trouble concentrating in the last two weeks, forced expiratory volume in one second (FEV1) z-score and having a job code or historical job code of a typist.

### Sensitivity analyses

Our meta-regression to identify the location of the best-fitting discontinuity (results shown in Table 1 and Table S3). This showed that the outcomes with the largest association with the origin in October were: age and year ended full time education, having O levels/GCSEs, having CSEs, having a college/university degree, comparative height at age 10, age starting smoking in former smokers, trouble concentrating in last two weeks, forced expiratory volume in one second (FEV1) Z-score, job code of typist, historical job code of typist, mean MO in tapetum on FA skeleton (right), mean OD in tapetum on FA skeleton (right) and mean MO in inferior cerebellar peduncle on FA skeleton (left).

## Discussion

In this study we conducted hypothesis-free IV-pheWAS to explore the effects of relative age in school year in around 383 000 UK Biobank participants born in England. We found 21 phenotypes that were associated with relative age, 13 of which displayed a discontinuity at the transition between school years. A number of our results are somewhat trivial, being directly due to the difference in participants age in their school year. This includes associations of younger relative age with being more likely to finish full-time education at an earlier age. It also includes an association between those with a younger relative age being more likely to report being shorter than their peers at age 10 (where participants likely think of their school year as their peers). Consequently, researchers using this UK Biobank variable in their analyses need to bear in mind that comparative height at age 10 is a somewhat biased measure of height in childhood. Furthermore, those with a younger relative age are more likely to start smoking at an earlier age (amongst former smokers). This is also somewhat expected given that peers are likely to start smoking at the same point in time, so those younger in the year will be younger when they start. Novel (non-trivial) results include those with a younger relative age being more likely to have better lung function in adulthood and having had trouble concentrating in the previous two weeks.

We found that younger relative age is associated with a lower likelihood of having educational qualifications, which is consistent with previous studies that report an association between younger relative age and lower educational attainment^1-7^. Several strategies have been proposed to address this inequality. Crawford, Dearden & Greaves (2013) found that children being different ages when they take national standardised tests is the main driver of relative age differences in educational attainment. They subsequently demonstrated that age-adjusting national achievement test scores (changing the number of points needed to achieve a certain grade depending on the age of the child, whilst keeping an absolute measure of children’s scores) effectively removes relative age differences, including the percentage of students reaching the government’s expected level^7^.

Looking at the persistence of relative age effects into adulthood,. in contrast to some previous studies^22-24^, but in line with others^7, 21, 26, 27^, we found little evidence of long term effects on employment prospects or earnings, with our only potential effect being having a job code or historical job code of a typist. However, this may be due to the extent and nature of employment variables in the UK Biobank. For example, analyses using individual occupation types will lack power. In addition, we did not find evidence of an effect of relative age on current employment status, but this may be due to the fact that around a third of UK Biobank participants are retired, so this variable is not necessarily a measure of occupational success. Nevertheless, a higher average total household income was strongly associated with being born in September vs August and week of birth, although it did not pass the Bonferroni threshold of either IV-pheWAS. Furthermore, in accordance with existing research^7, 19, 21^, we also found little evidence of long term effects on adult mental health, with mental health phenotypes that passed the Bonferroni threshold, such as sensitivity and mood swings, not displaying a discontinuity when plotted.

The little body of research previously conducted into the effect of relative age on adult physical health found no substantial differences in self-reported health^19^. In line with this, our IV-pheWAS only found one potential effect related to physical health. We found that adults with a younger relative age performed better in forced expiratory lung function tests. This result is supported by the fact that when we searched our results we also found that forced vital capacity z-score (another measure of lung function) was also positively associated with week of birth (*P*=5.97x10^−05^), meaning those with a younger relative age have a greater capacity. However, this did not pass the Bonferroni threshold. Our finding could possibly be the effect of children with a younger relative age being pushed in childhood to keep up with their bigger, older peers, with this process in effect training their lungs to give them better lung function as an adult. Alternative explanations include random chance or selection bias. This therefore warrants further exploration.

Our other potential effect of relative age on adult outcomes was that those with a younger relative age were also more likely to have had trouble concentrating in the last two weeks. Again, this could be a spurious finding, due to chance or selection. However, the fact that we also found having had “recent trouble concentrating on things” to be associated with relative age (albeit beyond the Bonferroni threshold) and that relative age has been associated with childhood diagnosis of attention-deficit/hyperactivity disorder^11-14^ suggests the effects of relative age on concentration in adulthood could warrant further investigation.

A key strength of our hypothesis-free IV-PheWAS approach is that it enables potentially novel effects to be uncovered. This possibility is amplified by the fact that the UK Biobank includes an extensive number and range of phenotypes, and that the PHESANT software allowed us to perform comprehensive phenome scans, rather than our set of outcomes being restricted to a homogenous subset^31^. We used the Bonferroni corrected *P* value threshold to reduce the likelihood of identifying effects that are due to chance. However, this method may be overly strict and therefore could have potentially resulted in our analyses missing some outcomes where relative age does have a true small effect. Hence, we have also reported the FDR threshold and provide the full set of results of our IV-pheWAS, so other researchers are able to follow-up other outcomes in future studies (see Table S1).

Some limitations of this work include the fact that the UK Biobank is not representative of the general UK population, with participants tending to be older, healthier, female and from less socioeconomically deprived areas^33^. If relative age in school year also affects participation in UK Biobank (which is likely given its suggested effect on educational attainment, which in turn is strongly associated with UK Biobank participation), then this could cause selection bias in estimating causal effects of relative age.

Further limitations include that PHESANT uses an automatic rule-based method to decide how each outcome is tested and it is possible that this may treat some variables inappropriately. In addition, models did not converge for a small number of traits so their relationship with relative age could not be reported. Lastly, because pheWAS aim to generate hypotheses, associations discovered should be replicated in an independent sample and using other study designs which have contrasting assumptions and biases^35^.

In summary, this study adds to the body of evidence that suggests that children born later in the academic year have lower educational attainment than those born earlier in the school year. Our results have implications for educational policy, as age-adjustment of national achievement test scores could potentially help to reduce these inequalities if presented alongside original scores. This could be particularly important for standardised national tests that determine children’s access to different types of schooling (Year Six SATs or Eleven Plus) or access to further education (GCSEs and A Levels). Future research should seek to replicate our identified effect on lung function in different populations, and investigate the mechanisms through which this effect may act. It could also further investigate the medium-term consequences of relative age on career prospects, and physical and mental health in early adulthood.

## Supporting information

Table S1 PHEWAS All Results

Table S2 PheWAS Outcomes without outputs

Table S3 Meta-regression with different origins results

## Data Availability

The UK Biobank dataset used to conduct the research in this paper is available via application directly to the UK Biobank. Applications are assessed for meeting the required criteria for access, including legal and ethics standards. More information regarding data access can be found at [https://www.ukbiobank.ac.uk/enable-your-research].

## Ethics approval

UK Biobank is approved by the National Health Service National Research Ethics Service (ref. 11/NW/0382; UK Biobank application number 16729).

## Supplementary data

Supplementary data are available at *IJE* online.

## Author contributions

K.T. initiated the study concept. M.A.dL., L.A.C.M. and N.M.D performed the statistical analysis. M.A.dL drafted the first version of the manuscript. All authors were involved in the interpretation of the data, critically reviewed and revised the manuscript, and approved the final version as submitted.

## Acknowledgements

This research has been conducted using the UK Biobank Resource under Application Number 16729. We thank Dr Beate Leppert for contributing to the early stages of this work. This work was carried out using the computational facilities of the Advanced Computing Research Centre, University of Bristol - http://www.bristol.ac.uk/acrc/.

