## Supplementary material for "Identifying potential effects of relative age in school year: an instrumental variable phenome-wide association study in the UK Biobank": Figures S1-20

**Figures S1-S2:** Forestplots for all outcomes that passed the Bonferroni threshold for IV^sep-aug^ or IV^weeks^ by month of birth vs September (IV^months^)

**Figure S1:** Age completed full time education by month of birth vs September

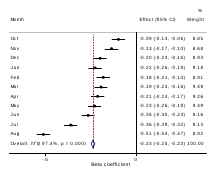

Note: Baseline (0) is month of September. Beta is the association between each month of birth and the outcome, relative to September.

**Figure S2:** Year ended full time education by month of birth vs September

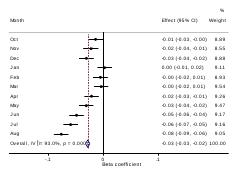

Note: Baseline (0) is month of September. Beta is the association between each month of birth and the outcome, relative to September.

**Figure S3:** Have CSEs or equivalent by month of birth vs September

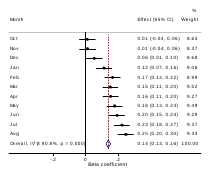

Note: Baseline (0) is month of September. Beta is the association between each month of birth and the outcome, relative to September.

**Figure S4:** Have O levels or GCSEs or equivalent by month of birth vs September

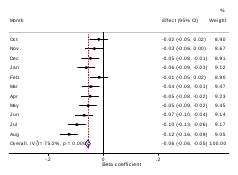

Note: Baseline (0) is month of September. Beta is the association between each month of birth and the outcome, relative to September.

**Figure S5:** Have A levels or AS levels or equivalent by month of birth vs September

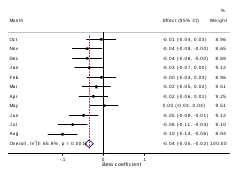

Note: Baseline (0) is month of September. Beta is the association between each month of birth and the outcome, relative to September.

**Figure S6:** Have college or university degree by month of birth vs September

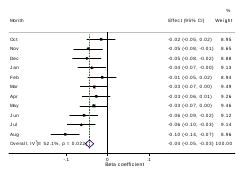

Note: Baseline (0) is month of September. Beta is the association between each month of birth and the outcome, relative to September.

**Figure S7:** Comparative height at age 10 by month of birth vs September

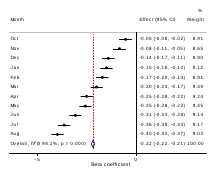

Note: Baseline (0) is month of September. Beta is the association between each month of birth and the outcome, relative to September.

**Figure S8:** Age started smoking in former smokers by month of birth vs September

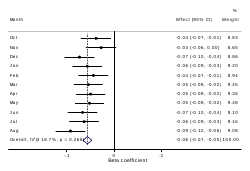

Note: Baseline (0) is month of September. Beta is the association between each month of birth and the outcome, relative to September.

**Figure S9:** Year job started by month of birth vs September

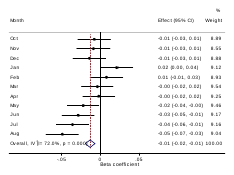

Note: Baseline (0) is month of September. Beta is the association between each month of birth and the outcome, relative to September.

**Figure S10:** Trouble concentrating on things in last 2 weeks by month of birth vs September

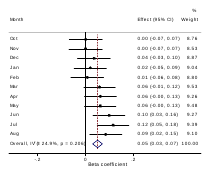

Note: Baseline (0) is month of September. Beta is the association between each month of birth and the outcome, relative to September.

**Figure S11:** Forced expiratory volume in 1-second (FEV1) Z-score by month of birth vs September

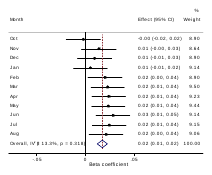

Note: Baseline (0) is month of September. Beta is the association between each month of birth and the outcome, relative to September.

**Figure S12:** Job code of typist or transcriber by month of birth vs September

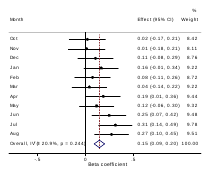

Note: Baseline (0) is month of September. Beta is the association between each month of birth and the outcome, relative to September.

**Figure S13:** Historical job code of typist by month of birth vs September

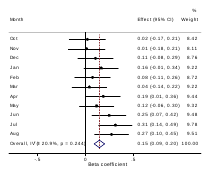

Note: Baseline (0) is month of September. Beta is the association between each month of birth and the outcome, relative to September.

**Figure S14:** Sensitivity/Feelings easily hurt by month of birth vs. September

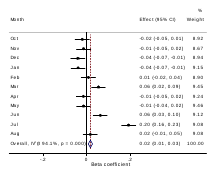

Note: Baseline (0) is month of September. Beta is the association between each month of birth and the outcome, relative to September.

**Figure S15:** Ever had bowel cancer screening by month of birth vs September

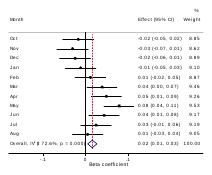

Note: Baseline (0) is month of September. Beta is the association between each month of birth and the outcome, relative to September.

**Figure S16:** Mood swings by month of birth vs September

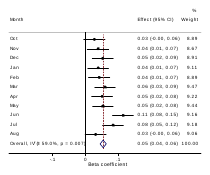

Note: Baseline (0) is month of September. Beta is the association between each month of birth and the outcome, relative to September.

**Figure S17:** 3mm asymmetry angle (right) by month of birth vs September

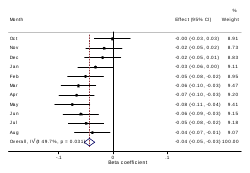

Note: Baseline (0) is month of September. Beta is the association between each month of birth and the outcome, relative to September.

**Figure S18:** Mean MO in tapetum on FA skeleton (right) by month of birth vs September

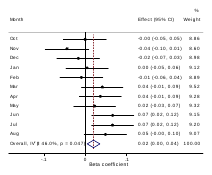

Note: Baseline (0) is month of September. Beta is the association between each month of birth and the outcome, relative to September.

**Figure S19:** Mean OD in tapetum on FA skeleton (right) by month of birth vs September

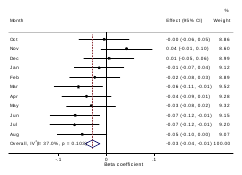

Note: Baseline (0) is month of September. Beta is the association between each month of birth and the outcome, relative to September.

**Figure 20:** Mean MO in inferior cerebellar peduncle on FA skeleton (left) by month of birth vs September

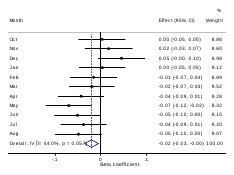

Note: Baseline (0) is month of September. Beta is the association between each month of birth and the outcome, relative to September.
